# Multimodal Deep Learning for Structural Heart Disease Prediction from ECG and Clinical Data

**DOI:** 10.64898/2026.02.22.26346793

**Authors:** Ajadi Nurudeen Ayobami, Afolabi Sulaiman Omotayo, Adenekan Ibrahim Oyeyemi, Jimoh Afeez Oyeshola, Ajayi Abayomi Olumuyiwa, Adeniran Teslim Ajibola, Adepoju Gboyega David, Hassan Nana Firdaous, Ajadi Salimot Aanu

## Abstract

This research presents multimodal deep learning for structural heart disease prediction. We evaluated multiple deep learning architectures, including TCN, Simple CNN, ResNet1d18, Light transformer and Hybrid model. The models were examined across the three seeds to ensure robustness, and bootstrap confidence interval is used to measure performance differences. TCN consistently outperforms other competing architectures, achieving statistically significant improvements with stable performance across runs. Similarly in predictive analysis, TCN has efficient computation and stable training compared to all competing architectures. Our results show that TCN emphasizes fairness evaluation when developing deep learning models for healthcare applications.

## 1.0 Introduction

Hypertension refers to the abnormalities of blood pressure. This is characterized by the continual pressure when the heart contract (systolic pressure) with a measure of at least 130mm Hg and the Diastolic pressure of at least 80mm Hg. Study shows that it affects approximately 116 million adults in the United States and more than 1 billion adults worldwide (Carey et al, 2022). The authors affirm that it is also related with high risk of cardiovascular disease (CVD) event, coronary heart disease, heart failure, stroke and death. According to the United States Centre for Disease Control (CDC, 2025), heart disease takes more lives than anything else in the United States.

Electrocardiograms (ECGs) and Echocardiograms (Echo) are cornerstones of cardiac testing. An ECG visualizes electrical activity and can be used to diagnose electrical rhythm issues or abnormal signals. An Echo uses real-time imaging to assess the heart’s functionality and internal anatomy. Both can assess and monitor the squeezing function and blood flow. These tools are reliable and provide the foundation for identifying potential heart problems.

Electrocardiography is among the most applied methods for diagnosing cardiovascular disorders because it is simple, less expensive, and non-invasive. An ECG measures the electric currents produced by the heart for each beat, which corresponds to the electric stimulation that makes the heart contract blood throughout the body (Liu et al., 2021). These electric currents travel through the body surface to spots where electrodes are placed, which in turn record them using an electrocardiograph (Liu et al., 2021). By interpreting the acquired records, cardiac abnormalities caused by irregular heartbeats or electric conduction can be estimated.

ECGs are employed in the diagnosis of conditions including arrhythmia, myocardial infarction, coronary heart disease, as well as cardiovascular complications in diabetes and hypertension (Liu et al., 2021). Apart from the diagnosis, it has been demonstrated that ECG abnormalities have prognostic uses in both short-term and long-term predictive values in cardiovascular diseases, including congestive heart failure and sudden cardiac death (Liu et al., 2021). This is particularly the case in the prognosis or analysis of electrical abnormalities on ECG since the abnormalities could remain symptom-free until serious or fatal complications occur.

Deep learning utilizes a multi-layered neural network. It has been successfully used to analyze many complicated medical data, like ECG signals and echocardiographic images (Elias et al., 2022). Echocardiography is particularly well-suited for deep learning applications because ultrasound imaging can capture many kinds of spatial and temporal data. Echocardiograms are often enough to give diagnostic information for a wide range of heart problems, from cardiomyopathies to valvular heart disease. This makes them perfect for automated analysis (Ghorbani et al., 2020).

Many people are still employing old-fashioned techniques in the interpretation of ECG and Echo, which are time-consuming and vary from one individual to another. Professionals in the medical field are required to interpret the intricate patterns in the waveforms and images. In fact, even the most experienced physician could overlook initial and a typical presentation of a disease (Velandia et al., 2025). The inherent variability in the process underscores the continued need to identify ways in which computational approaches could be used to assist in the process. Deep learning algorithms that have been trained well on large datasets are able to identify patterns in the data that the naked eye does not immediately recognize. Such algorithms have been shown to be equivalent to, if not better than, the expertise of the professional in cardiac medicine (Velandia et al., 2025).

The data from ECGs and ECHOs may be difficult to interpret manually since it is increasing and becoming more complex every day. Deep learning becomes a great approach to look for relevant data from images and signals not interpreted before. It is always possible to view data on a real-time scale. Deep learning makes it possible to identify hidden connections from ECG and ECHO data, which may be of great relevance to identify health concerns early, identify at-risk patients, and gain improved outcomes.

This research focuses on the application of deep learning algorithms to cardiovascular disease. Our main objectives are to (i) Construct 95% Bootstrap confidence interval across the three seeds (ii) Compare the efficiency of five (5) deep learning algorithms (iii) Apply fairness analysis to handle biasedness in the models (iv) Apply a masked-modelling approach in handling missing data in the EchoNext datasets (v) Evaluate the clinical deployment metrics of our models.

The remaining sections of this paper are arranged as follows: Section 2 reviews the application of Deep Learning techniques to Cardiovascular diseases. Section 3 presents the Deep Learning algorithms and the performance metrics used for the Analysis. Section 4 gives the data description, analysis and discusses the results. Section 5 presents the conclusion and recommendation of the research.

## 2.0 Literature Review

Deep learning has recently become an important tool for analyzing and interpreting electrocardiograms (ECG) and echocardiograms to help in disease detection, phenotyping, and the prediction of clinical outcomes (Ghorbani et al., 2020; Schots et al., 2025). Several neural network architectures form the foundation of medical deep learning. They include, Convolutional Neural Networks (CNNs) which is a kind of framework designed for recognizing patterns and extracting features from images and waveform data (Schots et al., 2025), Residual Networks (ResNets) are a specialized form of CNN architectures that improve information flow across deep networks (Schots et al., 2025). In optimization of sequential data analysis, Recurrent Neural Networks (RNNs) and Long Short-Term Memory (LSTM) have proven useful, because they can capture temporal dependencies in time-series signals like ECG recordings. Recent research has focused on hybrid model frameworks that combine these architectures. For instance, CNNs may be used for spatial or image-based feature extraction, while RNNs or LSTMs model temporal relationships, enabling the analysis of both spatial and temporal characteristics of physiological signals (Dhandapani et al., 2025). Across several cardiovascular applications, deep learning models have shown performance comparable to, and in some cases exceeding, that of medical experts. In the context of disease detection and prediction, DL models have achieved high accuracy in identifying pulmonary hypertension (PH) using ECG data alone, with reported AUC values of approximately 0.89. These models have also demonstrated the ability to predict PH up to two years before clinical diagnosis (Aras et al., 2023). In structural and functional diseases, DL approaches have been successfully applied to identify hypertrophic cardiomyopathy (HCM), valvular heart disease, and left ventricular dysfunction. The EchoNext model applied to echocardiograms can accurately estimate ejection fraction and identify local structures like pacemaker leads.

Acute and future events models like ECG-MACE utilize a multi-task learning approach to predict major adverse cardiovascular events (MACE), such as myocardial infarction, also known as heart attack, heart failure, and mortality (Lin et al., 2025). Furthermore, an AUROC of 0.90 has been reported for ECG-MACE in predicting hospitalization requiring heart failure within a one-year timeframe. When tested on a control subgroup, individuals with no history of major cardiovascular comorbidities such as hypertension or diabetes, the model’s AUROC for heart failure increased to 0.93. in comparison to other models, the multi-task approach utilized by ECG-MACE (0.90) demonstrated superior performance compared to a single-task learning model, which achieved an AUROC of only 0.83 for heart failure. Additionally, the model outperformed the Framingham risk score (FRS) in predicting heart failure at both 1-year and 5-year follow-up intervals. DL can predict phenotypes that are not readily apparent to human interpreters, such as a patient’s age, sex, height, and weight, directly from cardiac imaging. EchoNext model, has demonstrated that DL can predict these phenotypes directly from echocardiogram images. Models can estimate chronological age with an R^2^ of 0.46 and identify biological sex with an AUROC of 0.88. This has been mirrored in 12-lead ECG studies, which can also estimate “heart age,” a metric that provides additional information on cardiovascular risk beyond chronological age alone (Chang et al., 2022). DL models can predict a patient’s weight with R^2^ of 0.56, and height prediction with R^2^ of 0.33 by aggregating visual correlations across cardiac and extra-cardiac structures. Despite the advances, several challenges need to be solved before deep learning can be fully integrated into clinical practice. Some key concerns include patient safety, data quality, compliance with data protection regulations such as the General Data Protection Regulation (GDPR), limited model transparency, and difficulties integrating DL systems with electronic health records (EHRs). To address the hidden layers of deep learning, researchers use techniques that are interpretable such as Local Interpretable Model-Agnostic Explanations (LIME), saliency maps, and variational autoencoders (VAEs). These techniques help identify ECG segments or image regions that strongly influence model predictions, improving transparency and clinical trust (Ghorbani et al., 2020).

Research shows that echocardiography is very important during the diagnosis and treatment of different cardiovascular diseases, such as congenital heart diseases (Qi et al., 2024). Echocardiography interpretation for a cardiologist requires a diligent understanding and a high level of professional experience. Lack of technical know-how could result in misdiagnosis and can further cause a catastrophic issue. The author revealed that the application of AI and deep learning to the domain of Echocardiography is still at its early stage, with the capability to extract patterns (information) from large databases.

In addition, Krittanawong et al., (2023) explore the main potential of deep learning (DL) in addressing clinical informatics to real-world cardiology, especially via cardiovascular images with vast samples that demand expert skills. Generally, the author revealed that Deep Learning plays tremendous role in transforming large amounts of image data into cardiology insights and addressing different challenges in pandemic response. Moreover, (Ghorbani et al., 2020) introduce EchoNext which is a CNN based deep learning model that is trained on large amount of dataset, that can accurately and efficiently pinpoint cardia structure (such as pacemaker leads and enlarge left atrium), quantifying function and also forecast systematic traits such as sex, age and body metrics which outperform human limits. The strength of the model lies in the data-driven evidence, balanced metrics presentation, and validation through interpretation analysis, which builds trust by confirming the model’s focus on the relevant domain and flagging novel hypotheses.

## 3.0 Deep Learning Algorithms

In this research, we considered two families of algorithms including the CNN-family and the Transformer family. Three deep learning models (Simple CNN-1D, ResNet 1D-18 and TCN) under the CNN-family and two models (Light transformer and Hybrid) under the Transformer family were considered in our comparisons.

### 3.1 CNN Family

Convolutional neural networks (CNNs) are special-purpose neural network architecture for processing data with grid-like topology. Data with a grid topology includes time series data, which is like a one-dimensional grid with samples at regularly spaced intervals, and image data, which is like a two-dimensional grid of pixels. CNNs are neural networks that use convolution instead of regular matrix multiplication in at least one of their layers (Goodfellow et al, 2016).

#### 3.1.1 Simple CNN-1D

The Simple CNN-1D takes a 1-D input sequence and processes it through a stack of convolutional, activation, pooling, and fully connected layers to produce a final prediction (Goodfellow et al., 2016).

Let *x* ∈ ℝ^T^denotes a sequence of one-dimensional input signal, where *T* denotes length of the sequence. We defined the one-dimensional convolution with kernel

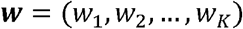

Producing the feature map

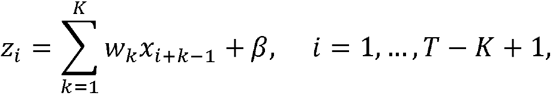

Where *β* is a bias parameter

This is followed by the application of a nonlinear activation function *g* (·), known as ReLU, which improves optimization and mitigates vanishing gradient issues.

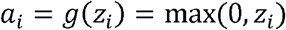

#### 3.1.2 ResNet 1D-18

Resnet 1D-18 is a one-dimensional version of the residual network architecture introduced to solve a key problem in deep learning, degradation problem, as networks become deeper, they become harder to train and may even perform worse leading to errors. In Resnet 1D-18, the architecture consists of an initial 1-D convolution followed by a sequence of residual blocks that together contain 18 weighted layers (He et al., 2016).

If *x* ∈ ℝ^T^denotes a one-dimensional input signal, a residual block computes the output

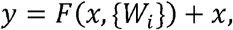

where *F* (*x*, {*Wi*})represents the residual function learned by the convolutional layers within the block.

For the standard two-layer block used in resnet-18, this residual function takes the form

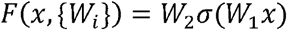

Where *σ* denotes the ReLU activation function. When the number of channels changes, the shortcut connection applies a learnable projection head to ensure dimensional compatibility. This resolves the optimization difficulties associated with increasing depth.

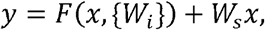

#### 3.1.3 TCN

Temporal Convolution Networks (TCNs) are convolution-based architecture introduced as simple and efficient alternatives to recurrent neural networks for sequence modelling tasks. **(Bai et al, 2018)**

For a one-dimensional input sequence *x* ∈ ℝ ^*n*^, a dilated convolution operation *y* is defined as

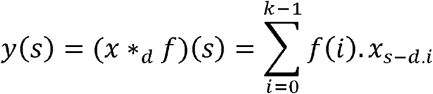

Where *k* is filter size, *d* is dilation factor and the subscript *s* − *d* · *i* only accesses past inputs.

As the dilation factor increases across layers, the network grows rapidly, allowing TCNs to model very long sequences using a relatively small number of layers. TCNs are constructed from residual blocks.

### 3.2 Transformer Family

Lightweight Transformer architectures combined with hybrid CNN–Transformer modules have been successfully applied to 3D medical image segmentation, as demonstrated by (Kuang et al., 2025). Although Transformers were originally introduced for natural language processing (NLP), where inputs are sequences of high dimensional word embeddings, similar principles apply to 1D encoded data. Language data shares structural similarities with image data when images are encoded as sequences of patches (Prince, 2023). This analogy enables Transformers to be adapted to vision tasks, where (CNNs) extract local spatial features effectively, and Transformers model long range dependencies and contextual relationships.

#### 3.2.1 Self Attention

Like the way human vision focuses on relevant details, a transformer model must emphasize important information, this mechanism is known as attention in transformer parlance.

If N inputs *x*_*n*_ form the columns of a D × N matrix **X**, the self attention computation is then:

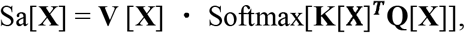

Where:

**V** is the value matrix computed as **V** [**X**] = **β**_v_ **1**^T^+ Ω_*V*_ **X, Q** is the query matrix computed as **Q**[**X**] = **β** q **1**^T^+ Ω_*q*_**X, K** is computed as Key matrix computed as **K**[**X**] = **β** _k_**1**^T^+ Ω_*k*_ **X**, the weight Ω∈ *R*^*D*X*D*^, the biases is defined as **β** ∈ *RD* and **1** is an N × 1 vector containing ones.

The SoftMax[·] takes a matrix and performs the SoftMax operation independently on each of its columns.

### 3.3 Evaluation Metrics

In this research, we evaluated our models by using relevant clinical metrics which are suitable in medical deep learning applications. These metrics are analyzed in this section.

#### 3.3.1 Sensitivity

Sensitivity can also be referred to as the True Positive Rate (TPR). It can be defined as the True Positives divided by the actual positives. This deals with the issue of how many of the actual Positive cases were captured and were labelled as True Positive by the model.

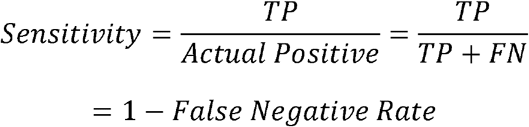

In the medical field, Recall (also called Sensitivity) is a measure of how many True Positive instances were found over total actual positives. From the perspective of the medical professional, a large Recall value suggests that there are few False Negatives, meaning that there are few actual situations that were not captured. In this case, Recall is telling the medical professional that a substantial number of relevant situations were correctly predicted. The consequences of a patient being misclassified as disease-free when, in fact, they are sick, is a large consequence of misclassification. They would miss out on critical medical care that would potentially save their life if they are suffering from a life-threatening illness.

#### 3.3.2 Specificity

Specificity measures how accurate a model is at identifying a negative. It refers to the number of true negative cases that are correctly identified and it is an indicator of how efficient a classifier is at identifying the negative cases. In the medical field, specificity is the measure of how many healthy individuals are accurately identified as not having the illness (i.e., the negative cases). The genuine negative rate is what is meant by specificity.

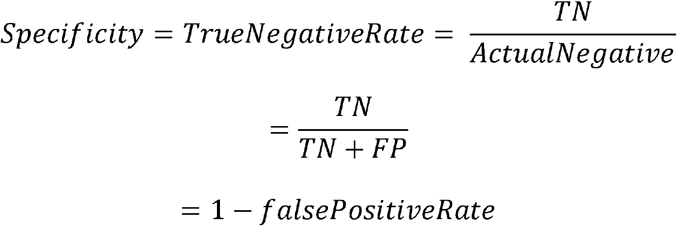

When we have great specificity, this means that false positive results happen very seldom. A poor specificity means that there are a lot of false positives.

#### 3.3.3 The area under the precision-recall curve (AUPRC)

The Area Under the Precision-Recall Curve (AUPRC) summarizes the different classification thresholds trade-off between precision and recall. The ROC curve considers true negatives, whereas the precision-recall curve focuses on performance for the positive class. Suitable for imbalanced datasets where the positive class is much smaller than the negative class, AUPRC is especially useful.

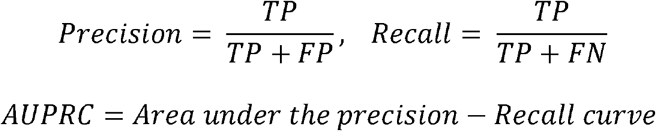

#### 3.3.4 Area Under the ROC Curve (AUC)

This assesses performance as a comparative trade-off between Sensitivity and Specificity. The Harmonic mean of the Sensitivity and Precision is the F-Measure. The model’s ability to make predictions gets better as the F-Measure increases.

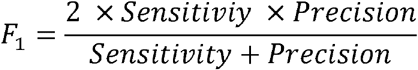

## 4.0 Data Analysis

In this study, we introduce a binary classification task to study Echo diagram confirmed structural heart disease (SHD) from electrocardiogram data (ECG) by using a variety of deep learning networks to train models for predicting SHD using 12-lead ECG waveform and Tabular clinical data.

### 4.1 Dataset Collection and Description

This research utilized the Echonext Mini-Model Dataset from PhysioNet (Elias and Finer 2025). The dataset contained 100,000 12-lead electrocardiograms (ECGs) with paired structural heart disease labels derived from echocardiography collected from the Columbia University Irving Medical Center. All these data were retrospectively collected from adult patients whose ages are at least 18 years. These datasets are collected within a one-year interval between 2008 and 2022. All ECGs data are paired with the corresponding metadata such as age, sex, heart rate, PR interval, QRS duration, QT interval and corrected QT interval and all these features were obtained from XML files obtained from GE MUSE Nx 10.2.

#### 4.1.1 Dataset Characteristics

The dataset used in this research is multimodal data which consists of the waveform modality-a 12-lead ECG signal sampled over 2500 time points and the tabular modality which is a fixed length vector of demographic and clinical attributes. The metadata and labels are loaded from a centralized CSV file while the waveform and tabular features are loaded from the NumPy arrays files as it has been preprocessed and partitioned into the training, validation, and test splits. The descriptive statistic for the dataset is presented in table 4.1.

**Table 4.1:** Descriptive Statistics of the datasets.

| Item | n | Mean | SD | Min | Max | Skew | Kurt | S.E |
| --- | --- | --- | --- | --- | --- | --- | --- | --- |
| age_at_ecg | 100000 | 61.17929 | 16.07706 | 18 | 90 | -0.44332 | -0.33246 | 0.05084 |
| acquisition_year | 100000 | 2016.122 | 3.857382 | 2008 | 2022 | -0.15195 | -1.06713 | 0.012198 |
| most_recent_ecg | 100000 | 0.36286 | 0.480827 | 0 | 1 | 0.570436 | -1.6746 | 0.001521 |
| ventricular_rate | 100000 | 83.57251 | 20.41984 | 2 | 223 | 0.828551 | 1.431224 | 0.064573 |
| atrial_rate | 99387 | 89.91937 | 43.92688 | 0 | 576 | 4.287425 | 23.75115 | 0.139337 |
| pr_interval | 89631 | 159.6174 | 32.52439 | 30 | 544 | 1.758367 | 8.073125 | 0.108638 |
| qrs_duration | 100000 | 94.71732 | 21.79784 | 4 | 254 | 1.504626 | 2.668122 | 0.068931 |
| qt_corrected | 99999 | 452.7469 | 39.99796 | 41 | 771 | 0.736756 | 4.209998 | 0.126485 |
| lvef_lte_45_flag | 100000 | 0.23892 | 0.426426 | 0 | 1 | 1.22451 | -0.50058 | 0.001348 |
| lvwt_gte_13_flag | 100000 | 0.2422 | 0.428417 | 0 | 1 | 1.203507 | -0.55157 | 0.001355 |
| aortic_stenosis | 100000 | 0.04054 | 0.197223 | 0 | 1 | 4.659318 | 19.70925 | 0.000624 |
| aortic_regurgitation | 100000 | 0.01264 | 0.111716 | 0 | 1 | 8.725063 | 74.12673 | 0.000353 |
| mitral_regurgitation | 100000 | 0.08451 | 0.278153 | 0 | 1 | 2.987512 | 6.92523 | 0.00088 |
| tricuspid_regurgitation | 100000 | 0.10651 | 0.308491 | 0 | 1 | 2.551078 | 4.507996 | 0.000976 |
| pulmonary_regurgitation | 100000 | 0.00821 | 0.090237 | 0 | 1 | 10.90004 | 116.811 | 0.000285 |
| rv_systolic_dysfunction | 100000 | 0.13243 | 0.338959 | 0 | 1 | 2.168825 | 2.703804 | 0.001072 |
| pericardial_effusion | 100000 | 0.03023 | 0.17122 | 0 | 1 | 5.487339 | 28.11089 | 0.000541 |
| pasp_gte_45_flag | 100000 | 0.18993 | 0.392248 | 0 | 1 | 1.580999 | 0.499559 | 0.00124 |
| tr_max_gte_32_flag | 100000 | 0.10212 | 0.302808 | 0 | 1 | 2.627953 | 4.906136 | 0.000958 |
| SHD | 100000 | 0.52188 | 0.499524 | 0 | 1 | -0.0876 | -1.99233 | 0.00158 |
| ivs_measurement | 90867 | 1.114982 | 0.258472 | 0.1 | 7.2 | 3.704675 | 61.44909 | 0.000857 |
| lvpw_measurement | 90875 | 1.062945 | 0.225891 | 0 | 6.8 | 2.197833 | 36.36205 | 0.000749 |
| pasp_value | 56576 | 41.54469 | 15.50229 | 10 | 184 | 1.582982 | 4.710942 | 0.065175 |
| tr_max_velocity_value | 45004 | 2.748951 | 0.608791 | 0.6 | 6.9 | 0.743793 | 1.638675 | 0.00287 |
| lvef_value | 91056 | 50.86804 | 15.00815 | 2.5 | 88.7 | -1.16097 | 0.359397 | 0.049736 |

#### 4.1.2 Data Augmentation and Mask Modelling

For effective deep neural networks (DNN) implementation, and to improve robustness, we applied ECG-specific augmentation to our dataset before being fed to the models. The use of this augmentation strategy is to reduce overfitting of the DNN models during training. Stochastic transformation like time shifting, amplitude scaling, additive gaussian noise and temporal masking are applied to the training data. These augmentations create a realistic simulation of random noise and Lead failure while preserving cardiac morphology to improve model generalizability and reliability.

Furthermore, our tabular data consists of missing values and labels that normally constitute ECG data in practice and as such we make use of mask-aware modelling (also refers to as missingness mask) so that the model understands when some clinical information is missing from the echocardiographic variables. For each such missing variable *j* in the metadata, a binary indicator is appended to the tabular feature vector before being to the model for the training.

#### 4.1.3 Descriptive Statistics

The descriptive statistics of the study in Table 4. 1 consist of 100,000 12-Lead ECGs, Echocardiographic and clinical information. The results show that majority of the patients who received ECGs are older individuals with an average age of 61.2 ±16.1 years. The average ventricular and atrial rates were 83.6 ± 20.4 20.4 beats per minute (bpm) and 89.9 ±43.9 bpm, respectively. This indicates the presence of an irregular distribution (right skew) with high volume and variability. The mean PR interval and QRS duration were 159.6 ±32.5 ms and 94.7 ±21.8 ms, respectively. The positive skewness indicated a minority of patients experienced conduction difficulties. The mean corrected QT interval (QTc) of 452.7 ± 40.0 ms indicates the entire sample’s QTc was a little above the normal range.

The results in Table 4.1 show multiple clinical manifestations of structural and functional heart disease. The results show that 23.9% of the group had a lower left ventricular ejection fraction (LVEF ≤ 45%) and 24.2% had left ventricular wall thickness (≥ 13 mm). 19.0% had pulmonary hypertension (pulmonary artery systolic pressure ≥ 45 mmHg). Valvular heart disease, which includes aortic, mitral, tricuspid, and pulmonary valve disorders, was less common overall, but had very uneven distributions; this is characteristic of a rare but significant condition. In 13.2% of cases, there was right ventricular systolic dysfunction, and pericardial effusion was observed in 3.0% of the cases.

The results of the echocardiography group revealed differences in the group as a whole. Septal and left ventricular posterior wall thicknesses were very right skewed, as would be expected in patients with left ventricular hypertrophy. The mean LVEF was 50.9 ± 15.0% and the distribution was left skewed. This demonstrates the low presence of left ventricular systolic dysfunction relative to the overall low mean. Both measured pulmonary pressure and tricuspid regurgitation velocity were widely distributed and non-normally distributed. Comprehensively, these results suggest a high and clinically diverse degree of cardiovascular disease in this group.

#### 4.1.4 Model Architecture

Since we are using a multimodal dataset, we constructed two different encoders network for better optimization of the dataset as shown in Figure 4.1. The ECG data are transformed and passed through the ECG encoders which consists of multiple ECG backbone architectures (Shallow 1D CNN, ResNet 1D, Temporal Convolution Network, Light Transformer Model, Hybrid Network) such that each backbone maps the ECG input to a fixed-dimensional latent embedding as depicted in Figure 4.1. In addition, tabular features are encoded using a multi-layer perceptron (MLP) with non-linear activations and dropout to improve generalization. The tabular encoder also produces a latent feature representation with the same dimensionality as the ECG embedding to allow for multimodal fusion.

**Figure 4.1:**
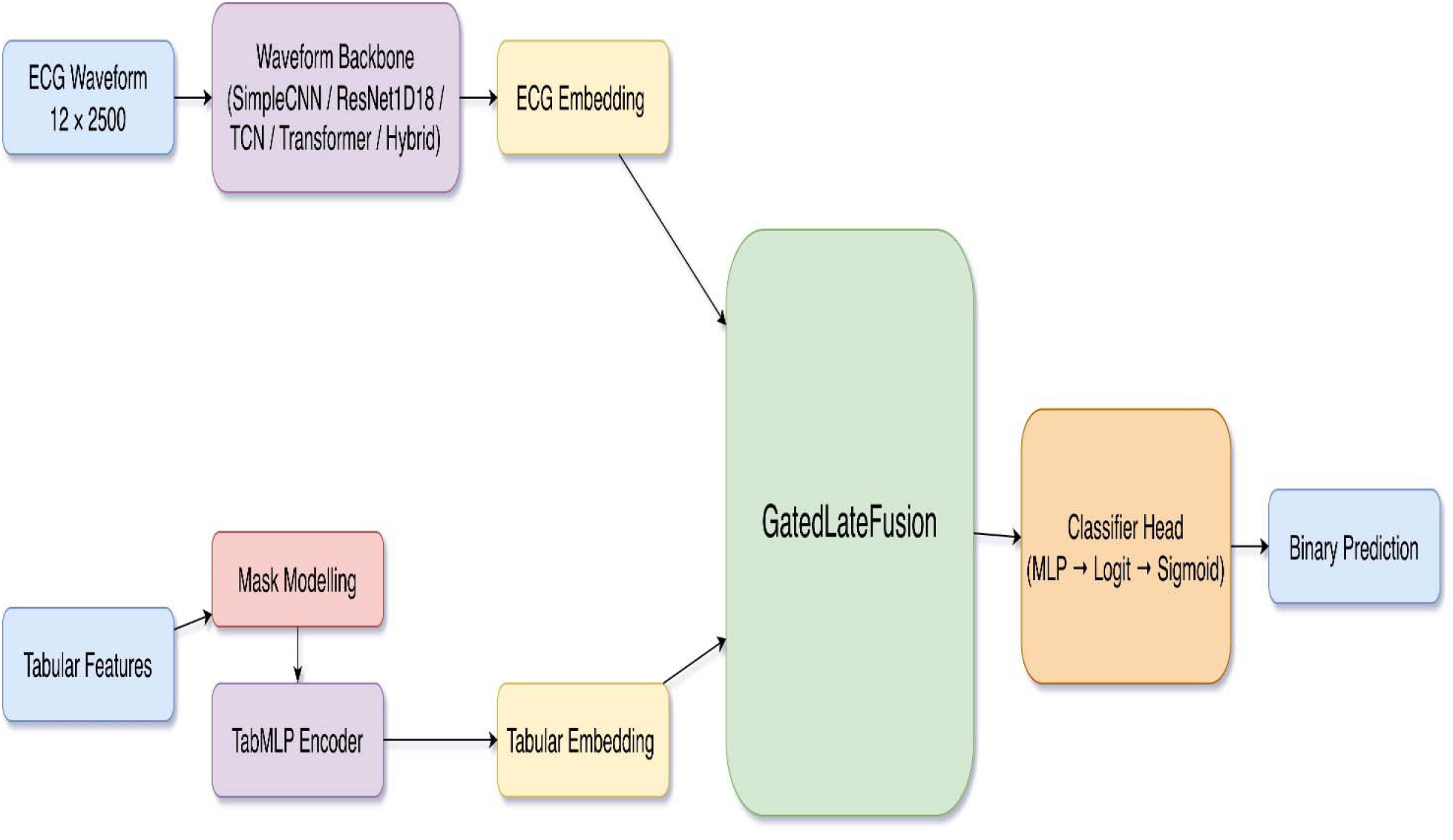
Model Architecture.

For the multimodal fusion strategy, we implemented an adaptive gated late-fusion in which the ECG and tabular features embeddings are first concatenated and pass through a gating network that outputs modality-specific weights. These weights are then normalized and fused to pass through the binary classification head to produce the final prediction.

### 4.2 Implementation Details

We started training all models from scratch with a batch size of 64 and learning rate of 3e-4 for learning consistency. For the loss function, we make use of binary cross entropy loss with logits and to address class imbalance, a positive class weighting factor is computed from the training data and incorporated into the loss function. We use *AdamW* as the optimizer with a weight decay value of 1e-4. A cosine learning rate scheduling with warm up is used to stabilize training in the early stages and improve model convergence. The training is set for 60 epochs with early stopping based on validation AUC and the mode checkpoint is selected based on highest validation AUC to prioritize ranking performance of the model rather than loss minimization only. For evaluation metrics, model performances are assessed using metrics like Area Under Receivers Operating Characteristic Curves (AUC), Area Under Precision Recall Curves (PR-AUC), Accuracy, Balanced Accuracy, Sensitivity, Specificity. To improve robustness of the model and reduce variance, each model is trained and evaluated using 3 independent random seeds and the result are reported as mean ± standard deviation across seeds. For fairness evaluation, we make use of equity-scaled AUC to assess performance of each model across two major sub-group (race/ethnicity, sex). All experiments are implemented with PyTorch on a Nvidia RTX A6000 with CUDA cores.

#### 4.2.1 Model Evaluation

In this study, we considered five different deep learning algorithms. Three of them are from the Convolutional Neural Network (CNN) family (the Simple CNN, the ResNet ID-18, and the Temporal Convolutional Network (TCN)), while the other two are from the Transformer family (the Light Transformer and the Hybrid). The hybrid algorithm integrates the CNN-Stem and Transformer encoders. The results in Table 4.2 are displayed as the mean ± Standard Deviation (SD). The mean represents the average performance, while the Standard Deviation provides insight on the variability of the data. Based on the AUC results, it appears that all Deep Learning (DL) algorithms perform comparably on all the classification tests. However, TCN appears to have the highest performance (0.8626 ± 0.0005). The Precision-Recall Area Under the Curve (PRAUC) result seems to be much more effective than the classification AUC in the case of unbalanced datasets. In this case, TCN performed best (0.8257 ± 0.0014). The result of the Accuracy (ACC) score seems to demonstrate that the TCN appears to be the best (0.7598±0.0036) across all competing models. The result of the F1 score appears to indicate that the best performance in balancing false positives and false negatives came from the Hybrid (0.7424±0.0041) and the Simple CNN (0.7408±0.0026). Given that the dataset had missing values, the metrics provided from Balanced Accuracy (BACC) in this case will represent the more reliable performance indicators of the data being unbalanced. The results indicate that the TCN has superior performance (0.7652±0.0023), followed by the Hybrid model (0.7662±0.0044). The Simple CNN (0.8374±0.0094) demonstrates great sensitivity, while the TCN model (0.7286±0.0181) exhibits better specificity performance. Our research focuses on AUC as our decision metric. Overall, TCN demonstrates superior performance across the evaluated measures.

**Table 4.2:**
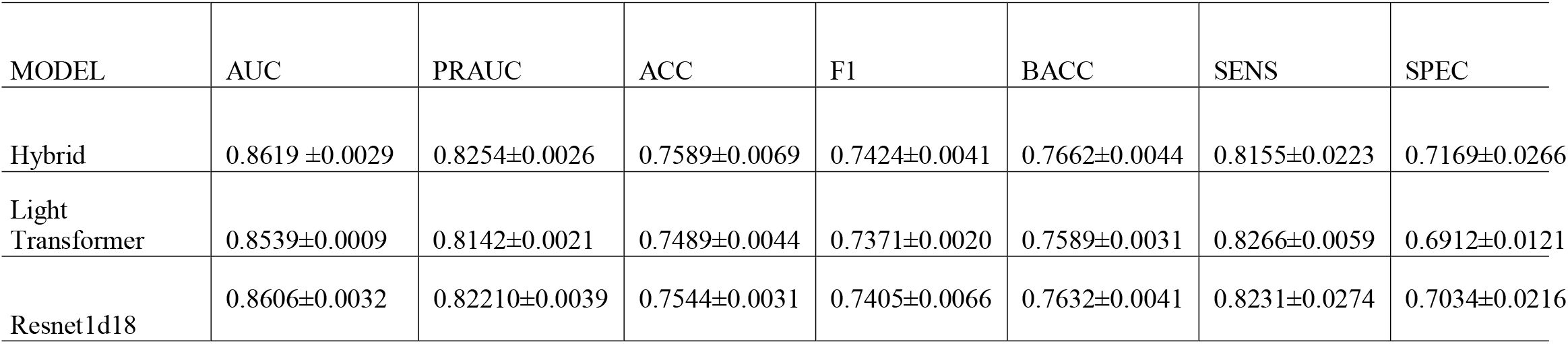
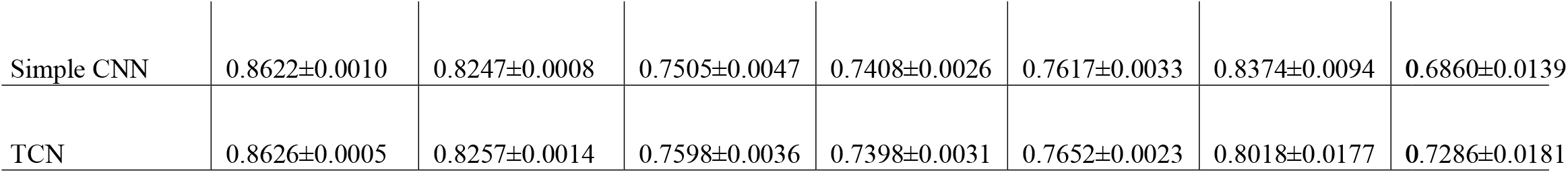
Evaluation Metrics for the Dataset.

#### 4.2.2 Fairness Metrics

The equity-scaled (Afolabi et al., 2025) AUC metrics from the model benchmark are shown in Figures 4.2 and 4.3. The overall result for the three seeds(1,2,3) are presented in Figure 4.4, but we highlighted the one that performed best. We considered the patients’ race/ethnicity and sex in the fairness analysis grouping. The fairness ranking for the racial group is presented in Figure 4.2. Among the subgroups, TCN has overall better performance on fairness in the race subgroups. Similarly, TCN exhibit overall better fairness performance in the Sex subgroup. Our results in Table 4.3 reflects that TCN and Hybrid models are fair with respect to both race and sex, which corresponds with the evaluation metrics.

**Table 4.3:**
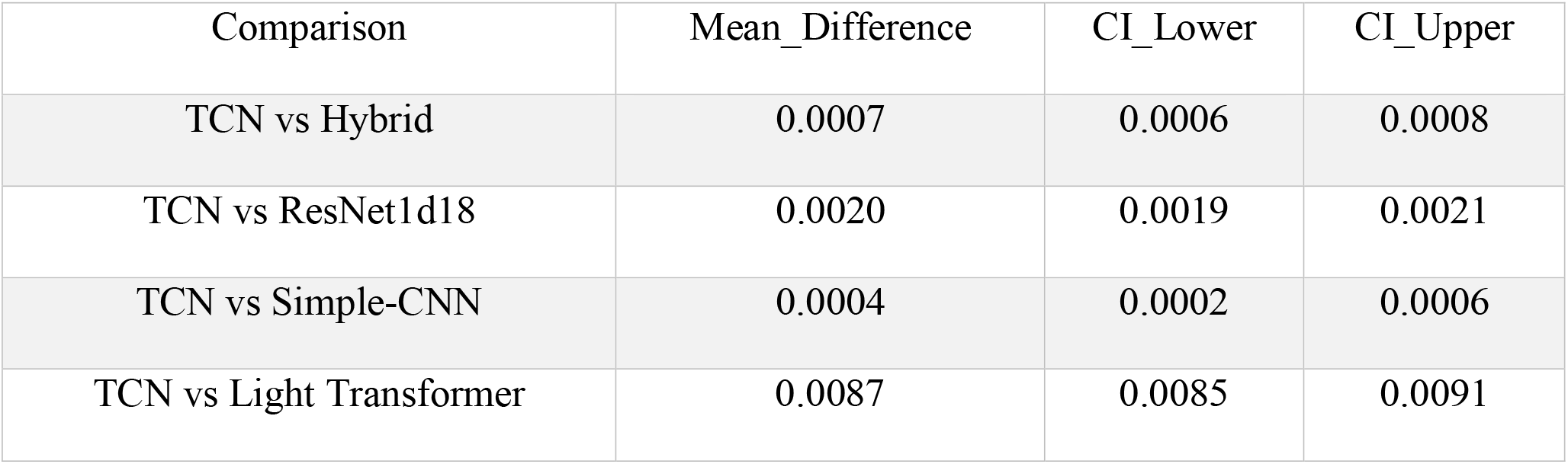
Bootstrap 95% Confidence Interval.

**Figure 4.2:**
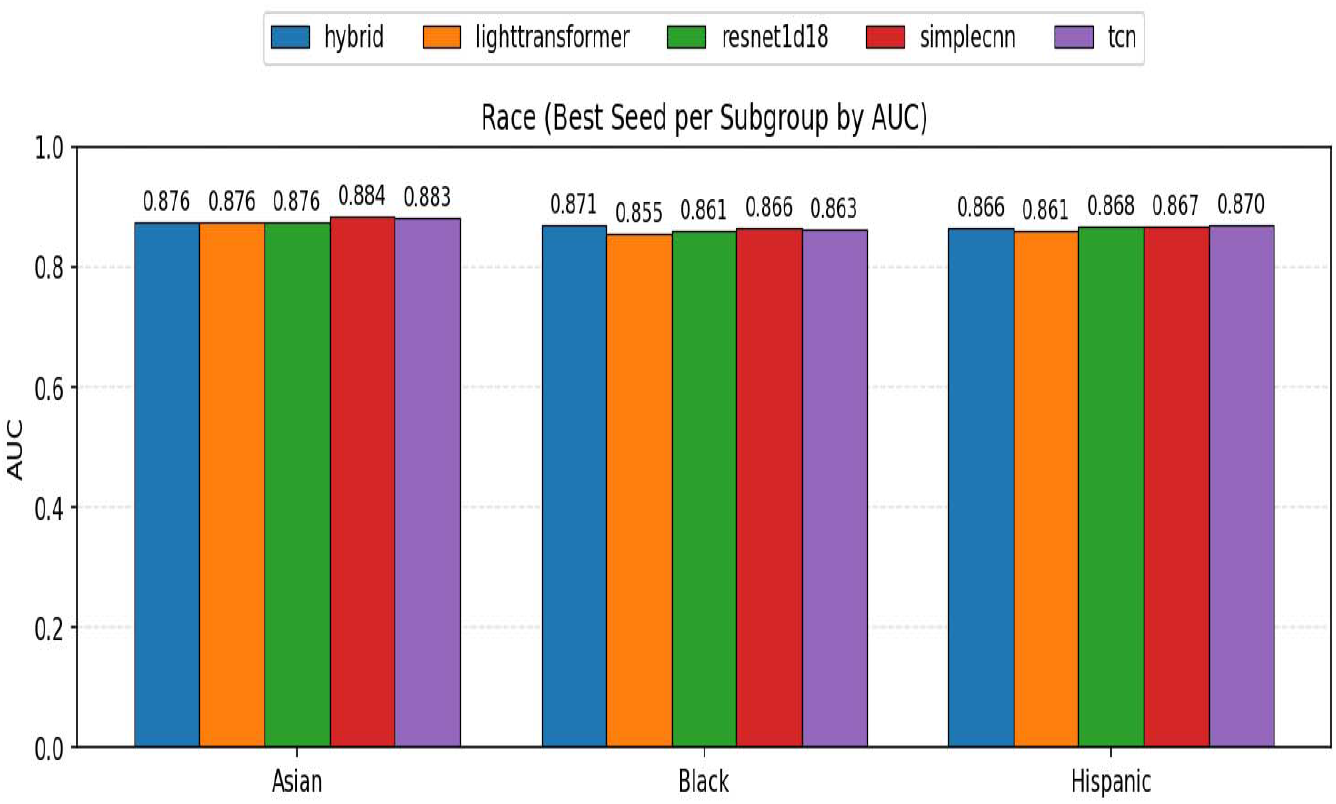
Fairness Metrics based on Race

**Figure 4.3:**
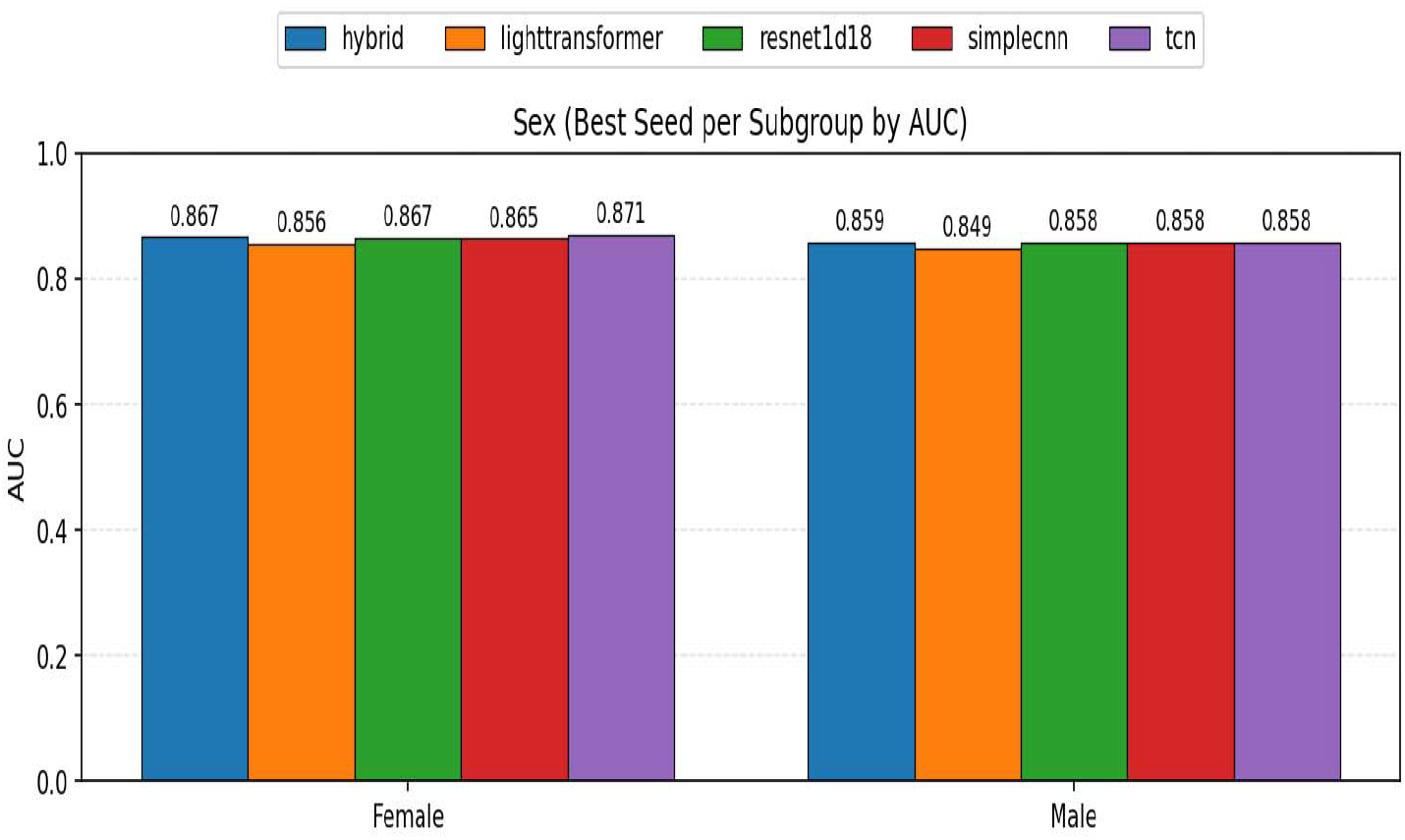
Fairness Metrics based on Sex

**Figure 4.4:**
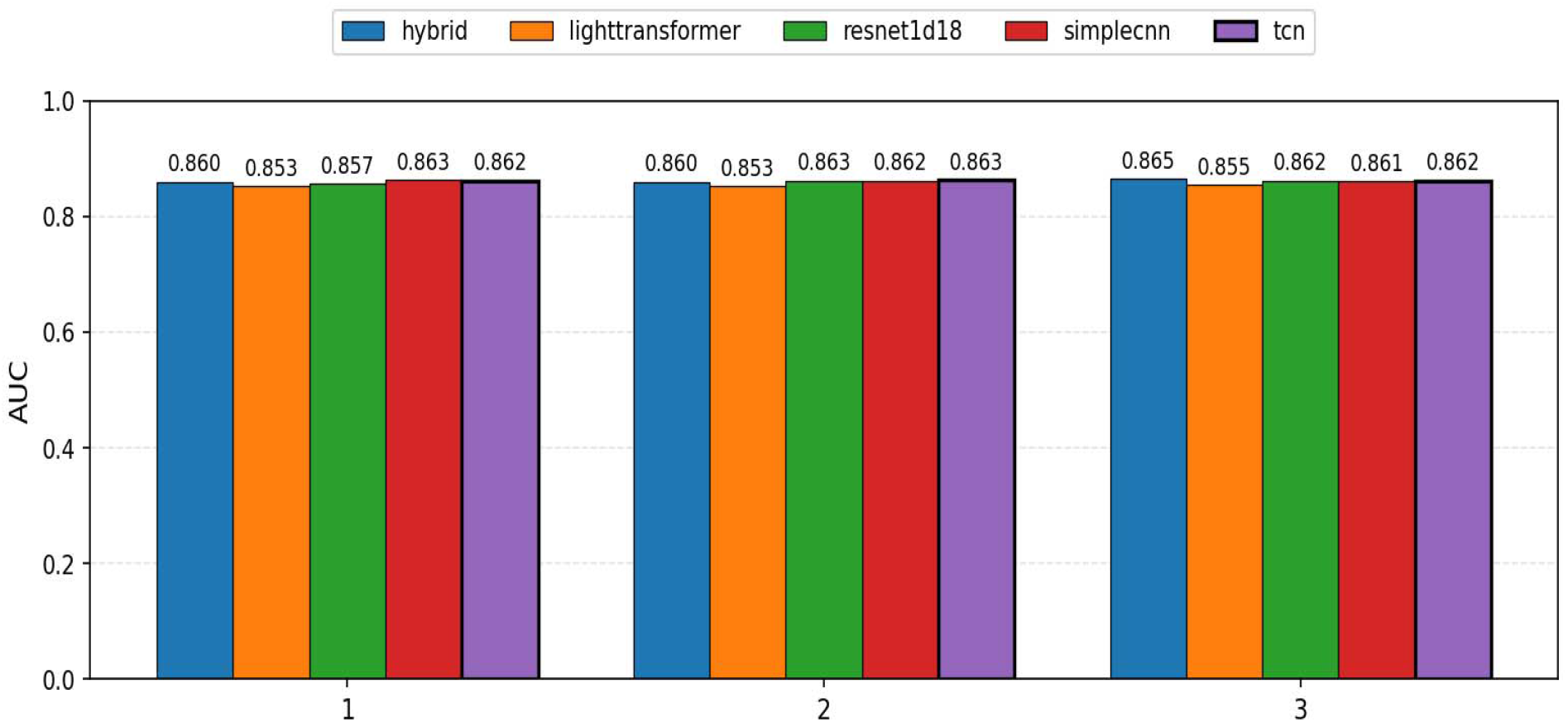
Fairness Metrics across the Three Seeds

#### 4.2.3 Bootstrapping Confidence Interval

The use of the bootstrapping confidence interval is one of our major contributions to knowledge. The result in table 4.2 has shown that TCN has the best performance, so we apply the bootstrapping method to validate our claim statistically. This method is based on resampling with replacement, and we generated 2000 simulations (B=2000). We presented the Bootstrap paired analysis across the three seeds in Table 4.3. The results demonstrated that the differences in AUC between the Temporal Convolutional Network (TCN), Hybrid, ResNet1D, and Simple CNN models were minimal. TCN showed a statistically significant improvement compared with the Hybrid, ResNet1d18 and Light Transformer models, as the confidence interval for the mean difference did not include zero. These findings are consistent with our results in Table 4.2

#### 4.2.3 Global SHAP Analysis

The physiological interpretation of the 12 leads ECG is presented in table 4.4 and the analysis of lead importance based on SHAP values in Figure 4.5 has shown that among the precordial leads, V1, V2 and V3 have the greatest impact on the prediction of structural heart disease. This is clinically reasonable, as these anterior and septal leads sense electrical signals from the right ventricle, the interventricular septum, and the anterior wall of the left ventricle, which are the areas that most frequently involve structural cardiac diseases such as ventricular hypertrophy, cardiomyopathies, and conduction disorders. Substantial changes in the myocardium structure also lead to changes in the depolarization and repolarization cycles that are not easy to recognize in a regular ECG and can be only found more effectively by deep learning models which is based on waveform analysis. The less significant reported for the lateral and inferior leads is indicative of the fact that the unique electrical activity of the diseased ventricles is more important in this cohort for disease prediction. Finally, the cardiological relevance of the most important leads identified by the model further reinforces the novel approach and prioritizes its potential as non-invasive screening method for early structural heart disease detection.

**Table 4.4:** Physiological Interpretation of the 12-Lead ECG.

| **Lead** | **Full Meaning** | **View** | **Main Cardiac Region** |
| --- | --- | --- | --- |
| Lead I | Right arm → Left arm | Left lateral view | High lateral wall of left ventricle |
| Lead II | Right arm → Left leg | Inferior view | Inferior wall of left ventricle |
| Lead III | Left arm → Left leg | Inferior view | Inferior wall of left ventricle |
| aVR | Augmented Voltage Right arm | Right upper view | Opposite global vector (usually negative) |
| aVL | Augmented Voltage Left arm | High lateral view | High lateral wall of left ventricle |
| aVF | Augmented Voltage Foot (left leg) | Inferior view | Inferior wall of left ventricle |
| V1 | 4th intercostal space, right sternal border | RV + septal view | Right ventricle and interventricular septum |
| V2 | 4th intercostal space, left sternal border | Septal view | Interventricular septum |
| V3 | Between V2 and V4 | Anterior view | Anterior wall / transition zone |
| V4 | 5th intercostal space, mid-clavicular line | Anterior view | Anterior wall of left ventricle |
| V5 | Left anterior axillary line | Lateral view | Lateral wall of left ventricle |
| V6 | Mid-axillary line | Lateral view | Lateral wall of left ventricle |

**Figure 4.5:**
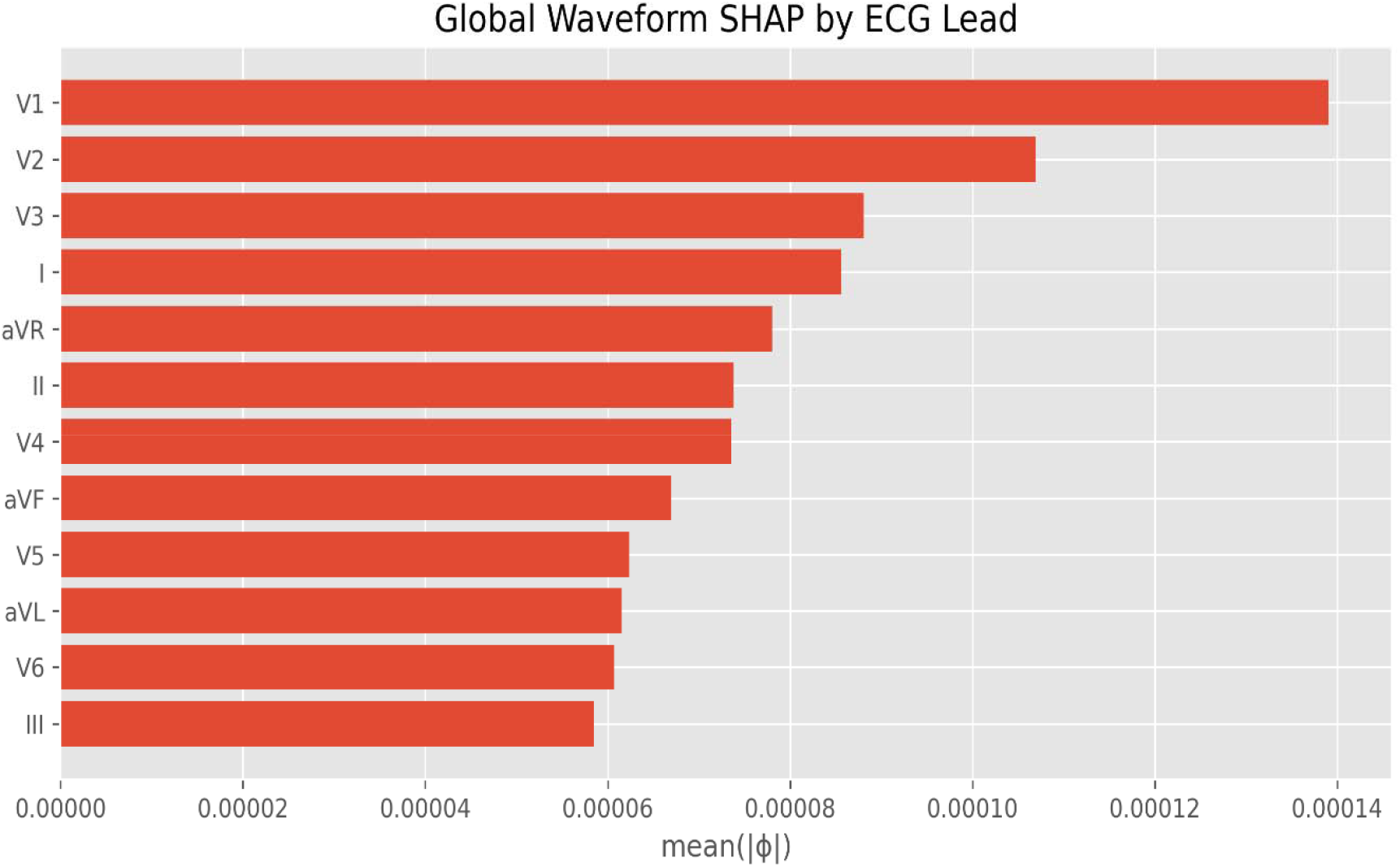
Global Waveform SHAP by ECG Lead

The Global SHAP analysis of the ECG waveform over time is presented in Figure 4.6. This method was adopted by (Afolabi et al., 2025) in detecing the significant feautures to the medical data set. The Figure shows how certain time spans of the ECG signal are useful in the predictive analysis of structural heart disease. The most SHAP values across the sensors stay at the same level determined the same amount of time. This means the same amount of sensor data integrity is equally tarnished. Multiple peaks indicate ample available/accessible data at a sensor time period, specifically during the most physiologically significant parts of the ECG and the most consider the QRS complex. These time stamps demonstrate the cycling of the heart; the process of ventricular systole and diastole, the processes that most structural heart disease problems involve, e.g. ventricular hypertrophy, cardiomyopathy, and conduction disturbances. The less SHAP values at the start and the end of the time stamps could be due to the beginning and end of the inner workings of the machine that produced the signal. Overall, the pattern presents the final biological and functional evidence of the efficacy and the reliability of the model for the potential examination of structural heart disease

**Figure 4.6:**
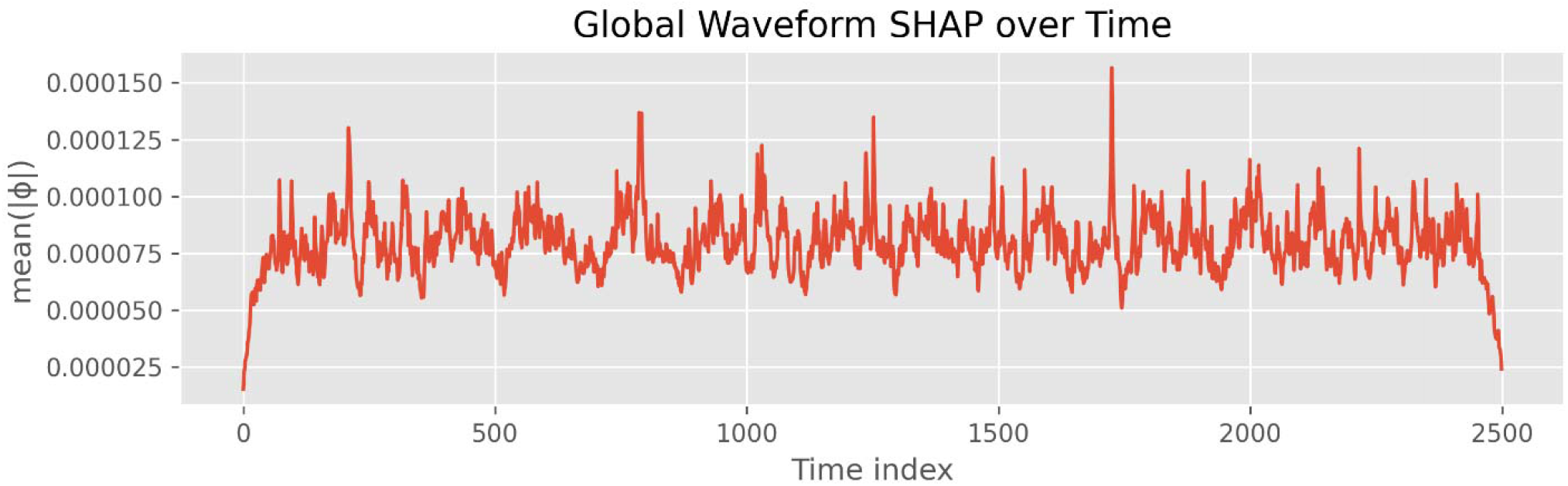
Global Waveform SHAP

## 5.0 Discussion and Conclusion

In this study, we utilized the EchoNext dataset from the Physionet database which is maintained and funded by the MIT laboratory of Computational physiology and supported by the National Institute of Biomedical Imaging and Bioengineering, and the National Heart Lung and Blood Institute and National Institute of Health. The aim of this study is to analyze various factors and clinical attributes that are vital in the detection of structural heart disease (SHD) using various deep neural network models for early and accurate diagnosis of impending heart failure and related disease.

Due to the multimodal nature of the EchoNext dataset which consists of clinical features, waveform feature and the patients’ demographic attributes, we understand the need for accurate modelling and scrutiny required in DNN training. Thus, we choose compatible and most-efficient DNN models for training, validating, and testing of the dataset. To enhance our analysis in medical dataset, the choice of validation metrics cannot be over emphasized. 1D image data in form of waveform features, necessitated the need for AUC metrics which is more resilient and preferable to accuracy. Also, for clinical interpretability, we utilized clinical metrics such as, Sensitivity, Specificity and PRAUC.

In addition, we presented the five model performances in terms of clinical metrics used to evaluate and compare individual models having trained, validated, and tested using three different seeds. Meanwhile, we also evaluated these models using bootstrapping confidence intervals for statistical significance. Since our dataset has demographic attributes, we also introduced fairness analysis by using equity-scaled AUC which penalized the model based on gender and race. This is to ensure the removal of biasedness in DNN models and its generalizability.

In overall, the temporal convolution network (TCN) achieves the best performance in terms of clinical interpretability metrics as well as the overall fairest models amongst the five different models evaluated in this research. We conclude that for best diagnostic modelling of SHD with this dataset using our methodology, the most reliable DNN model is the TCN as it has the highest AUC value and the fairest models amongst the competing models.

### 5.1 Recommendation and Further Research

Our aim is to make immense contribution to the continuous research and providing solution to sudden death which is prevalent among patient with structural heart diseases. This study recommends the use of TCN architecture for predicting the potential occurrence of heart diseases based on ECG and clinical information. Though the use of artificial intelligence in healthcare is yet to gain global acceptance by health practitioners, we believe that use of these innovative ideas will assist in reducing the risk of structural heart disease. This research is limited to 100,000 datasets; however future research can adopt the utilization of more datasets for robust and generalizability application and usage.

## Data Availability

All data produced are available online at https://physionet.org/content/echonext/1.1.0/

